# A Mass Casualty Event in a Gaza War Hospital

**DOI:** 10.1101/2024.11.14.24316980

**Authors:** Abdalkarim Alsalqawi, Khader Abu Tahoon, Adnan Nijim, Mohammed Halimy, Richard Villar

## Abstract

This study examines the impact of a mass casualty event (MCE) on a Gaza hospital during a military operation in June 2024, highlighting unique challenges encountered in conflict-zone medical care. The research analyses 47 casualties received by the Shuhada al-Aqsa Hospital in Deir-al-Balah, Gaza, over a concentrated period. Data collected include demographics, injury types, and pre-hospital and emergency department treatment delays. Findings reveal a predominance of blast injuries (76.6%), with compound injuries being present in 78.7% of casualties, resulting in a high injury burden (mean 1.68 injuries per casualty). The mean age of all casualties was 30.57 years. Mortality reached 14.89%, with females disproportionately affected, comprising 71.43% of fatalities. Transportation delays averaged 18.7 minutes, often without pre-arrival notification, underscoring logistical challenges in the Gaza conflict setting. In-hospital delays for treatment averaged 17.4 minutes. Gender disparities in trauma outcomes suggest biological and sociocultural factors influencing survival rates. The study emphasises the need for conflict-sensitive, gender-informed medical interventions, improved trauma care resources, and pre-hospital coordination protocols. This work aims to inform international medical volunteers on the realities of conflict-zone healthcare and support system-level improvements in MCE responses in resource-limited settings.

## Introduction

Mass casualty events (MCEs) are situations where the number of casualties overwhelms the available medical resources, necessitating the prioritisation of care through triage and the efficient use of available resources to save as many lives as possible^i^. MCEs can arise from various circumstances, including natural disasters, industrial accidents, terrorist attacks, and military conflicts.

In developed countries, MCEs are typically associated with events such as mass shootings, large-scale traffic accidents, or terrorist bombings. For instance, the 2015 Paris attacks resulted in 130 deaths and 416 injuries, almost 100 critically^ii^. Hospitals in the region rapidly mobilised to treat patients with a range of injuries, including gunshot wounds and blast injuries. The overall mortality for those who reached the hospital was less than 2%^iii^, attributed to the immediate availability of advanced trauma care and the implementation of well-coordinated emergency response protocols.

Similarly, the Boston Marathon bombing in 2013, which resulted in three people being killed and hundreds injured, including 17 who lost their limbs^iv^, demonstrated the esectiveness of pre-established mass casualty protocols. Hospitals in Boston reported no in-hospital deaths^v^. The injuries included shrapnel wounds, traumatic amputations, and blast-related concussions, but the swift triage and access to surgical interventions were key factors in the high survival rate.

In contrast, MCEs in conflict zones such as Gaza present unique challenges that are not typically encountered in non-conflict zone settings. The Gaza Strip, an area with a dense population and limited medical infrastructure, has been subjected to recurrent military conflicts, resulting in frequent MCEs characterised by a high volume of explosive injuries and severe trauma. The ongoing nature of the conflict exacerbates the strain on medical resources, making it difficult for hospitals to manage the influx of patients effectively. In such settings, healthcare providers often face shortages of essential medical supplies, limited access to advanced medical technologies, and overwhelming patient volumes that significantly impact survival outcomes.

Given these differences, it is crucial to understand how the management of MCEs in a region such as Gaza compares with MCEs in developed countries where conflict is not occurring. This understanding is particularly important for humanitarian medical staff from non-conflict zones who may volunteer to work in Gaza. These individuals are often accustomed to working in well-equipped healthcare systems with ample resources and may find the conditions in Gaza challenging. Understanding the injury patterns, resource limitations, and survival outcomes in a Gaza hospital during an MCE can help these volunteers prepare for the realities of working in a conflict zone.

### Background

On 8 June 2024, at around the middle of the day, the Israeli military attempted a hostage rescue operation in the UNRWA Nuseirat refugee camp of central Gaza. The operation’s objective was to free multiple hostages taken during the 7 October 2023 attack on Israel. Unanticipated operational events led to what was described as a “crazy bombardment”^vi^, which destroyed apartment buildings and even entire residential blocks throughout the camp. Available vehicles and ambulances rushed wounded people to the Shuhada al-Aqsa Hospital, three kilometres away, for treatment. The total number of casualties is disputed, with Israeli and Palestinian totals differing drastically. The Gaza Health Ministry and local health officials stated at least 274 Palestinians were killed and around 700 wounded^vii^. The Israel version differs with fewer than 100 Palestinians said to have been killed during the operation.

Irrespective of the disagreement in respect of the number of casualties, and how the military operation was conducted, an MCE occurred at the Shuhada al-Aqsa Hospital. Approximately 200 patients were admitted to the Emergency Department (ED) of the hospital because of this military activity, which has been named a massacre^viii^.

Because MCEs are situations where the number of casualties overwhelms the available medical resources, during the Israeli-Palestinian conflict that began in 2023 every day saw a MCE in one or more locations in Gaza. This study is a snapshot view of one of them. Our purpose is to demonstrate what can be expected of an MCE during an ongoing conflict in a resource-deprived setting, any lessons learned, and to compare it with what might be expected during an MCE in a more developed part of the world.

## Materials and Methods

We here describe 47 patients who presented to the 16-berthed ED of the Shuhada al-Aqsa Hospital from 1125-1340hrs on 8 June 2024, as a snapshot view of an MCE in progress. The MCE lasted for a longer period than this (8.5 hours), involved a total of approximately 200 patients presenting to the ED, and was regarded as finished by 2000 hrs that day. These 47 patients thus represent 23.5% of the total.

We obtained the following information for each casualty in the ED during this period:

1. Gender
2. Age
3. Diagnosis
4. Compound or closed injury
5. Cause of injury (explosion, gunshot, other)
6. Nature of injury
7. Time of injury
8. Time of arrival at ED
9. Time when casualty first treated in ED
10. Number of injuries sustained
11. Mode of transport to ED
12. Notice (or no notice) given of patient’s arrival at ED
13. Length of stay (hours) in ED
14. To where discharged from ED
15. Whether or not the casualty died in ED

### Permission

Permission to undertake this study, and report these findings, was given by the administration and Medical Director of Shuhada al-Aqsa Hospital, Deir-al-Balah, Gaza. We have ensured that no patient is identifiable in this study. Because of ongoing conflict, there was no Ethics Board in existence, but we considered this to be unnecessary in any event.

## Results

Of the 47 casualties, 23 (48.94%) were male and 24 (51.06%) were female. The mean age of all casualties was 30.57 years (6 to 70), with a mean for males of 30.89 years (9 to 60) and for females of 31.46 years (6 to 70). The age distribution of the casualties may be seen in Table 1.

**Table 1.** age distribution of the 47 casualties.

| Age range (years) | Number (%) |
| --- | --- |
| All ages | 47 (100%) |
| <18 | 7 (14.89%) |
| 18-35 | 26 (55.32%) |
| >35 | 14 (29.79%) |

For the 47 casualties, there were 79 injuries, distributed almost equally between males (n=39) and females (n=40). There were thus 1.68 injuries/casualty (79/47=1.68). The age distribution of these injuries may be seen in Table 2 and the nature of the injuries can be seen in Table 3.

**Table 2.** age distribution of the 79 injuries in the 47 casualties.

| Age range (years) | Number of injuries (%) | Number of injuries/casualty |
| --- | --- | --- |
| All ages | 79 (100%) | 1.68 |
| <18 | 8 (10.13%) | 1.14 |
| 18-35 | 48 (60.76%) | 1.85 |
| >35 | 23 (29.11%) | 1.64 |

**Table 3.** nature of the 79 injuries in the 47 casualties.

| Injury | Number |
| --- | --- |
| Rectal injury | 4 |
| Sigmoid injury | 1 |
| Subarachnoid haemorrhage | 1 |
| Small bowel injury | 3 |
| Cardiac tamponade | 1 |
| Humeral fracture | 5 |
| Tibial fracture | 4 |
| Pneumothorax | 4 |
| Hip disarticulation | 1 |
| Vertebral fracture | 1 |
| Lower limb amputation | 2 |
| Radial and/or ulnar fracture | 7 |
| Burns | 4 |
| Proximal femoral fracture | 1 |
| Pelvic fracture | 5 |
| Stomach injury | 2 |
| Liver injury | 2 |
| Subgaleal haematoma | 1 |
| Skin loss | 4 |
| Laceration | 2 |
| Upper limb amputation | 1 |
| Haemothorax | 3 |
| Fibular fracture | 2 |
| Femoral shaft fracture | 1 |
| Metacarpal fracture | 2 |
| Radial artery injury | 1 |
| Skull fracture | 1 |
| Metatarsal fracture | 2 |
| Brain leakage | 1 |
| Eye globe rupture | 1 |
| Neck injury | 1 |
| Spleen injury | 1 |
| Diaphragm injury | 1 |
| Spinal cord injury | 2 |
| Kidney injury | 1 |
| External iliac artery injury | 1 |
| Femoral vessel injury | 1 |
| Tendon injury | 1 |

For the 47 casualties with their 79 injuries, 37 (78.7%) had a compound (open) injury and 10 (21.3%) did not. The age distribution of these injuries may be seen in Table 4.

**Table 4.** age distribution for those with a compound injury.

| **Age range (years)** | **Number (%) of casualties with a compound injury** | **Number (%) of casualties without a compound injury** |
| --- | --- | --- |
| All ages | 37 (78.72%) | 10 (21.28%) |
| <18 | 4 (57.14%) | 3 (42.86%) |
| 18-35 | 23 (88.46%) | 3 (11.54%) |
| >35 | 10 (71.42%) | 4 (28.58%) |

Of the 47 casualties, 36 (76.60%) were injured by an explosion, 7 (14.89%) by a gunshot wound (GSW), 3 (6.38%) by a fall, and 1 (2.13%) in a road traffic accident (RTA). Thirty-six (76.6%) of the casualties were transported to hospital by ambulance and 11 (23.4%) by private car. Advance notice was given to the hospital of a casualty’s arrival in only 8 (17.02%) cases.

The mean delay (range) for the transportation of a casualty from their Point of Injury (POI) to the hospital was 18.7 (8 to 35) minutes, the mean for males being 20.04 (8 to 35) minutes and the mean for females being 17.42 (10 to 30) minutes. There was little difference in transportation delay between the age groups, as demonstrated in Table 5.

**Table 5.** transportation delays from injury to hospital in the different age groups.

| **Age range (years)** | **Mean (range) transportation delay (minutes)** |
| --- | --- |
| All ages | 18.7 (8 to 35) |
| <18 | 18.57 (10 to 35) |
| 18-35 | 18.62 (8 to 30) |
| >35 | 18.93 (10 to 30) |

The mean delay (range) in receiving treatment once in the ED of the hospital was 17.4 (5 to 45) minutes for all casualties, 17.36 (5 to 45) minutes for males and 17.13 (5 to 45) minutes for females. There was little difference in treatment delay between the age groups, as demonstrated in Table 6

**Table 6.** delay in receiving treatment once in the Emergency Room (ER)

| **Age range (years)** | **Delay (range) in minutes** |
| --- | --- |
| All ages | 17.4 (5 to 45) |
| <18 | 18.06 (10 to 45) |
| 18-35 | 17.36 (5 to 40) |
| >35 | 17.40 (5 to 45) |

Of the 47 casualties, 7 (14.89%) died in the ED. The mean age of those who died was 32.43 (17 to 70) years. Two (28.57%) were male and 5 (71.43%) were female. All seven (100%) casualties, who eventually died in the ED had been transported from their POI to hospital by ambulance. No notice had been given to the hospital of their arrival. For those who died, the mean number of injuries for male victims was 1.0, but 1.8 injuries for female victims. For the seven victims, four died from haemorrhage, one from septic shock, while the cause of death for two victims was not recorded. These figures suggest that 8.69% of male casualties perished but 20.83% of female casualties also perished. All who died were victims of explosive blast and not of GSW.

As 7 of the 47 casualties perished in the ED, there were 40 survivors. The mean length of stay in the ED for these 40 individuals was 21.58 (2 to 96) hours. This length of stay varied widely, depending upon the age range, as shown in Table 7.

**Table 7.** length of stay in ED (hours) by age range for survivors (n=40)

| **Age range (years)** | **Length of stay in the ED (hours)** |
| --- | --- |
| All ages | 21.58 (2 to 96) |
| <18 | 31.17 (3 to 96) |
| 18-35 | 20.32 (3 to 72) |
| >35 | 19.08 (2 to 72) |

For the survivors, their location of discharge from the ED was the ward (n=28, 70%), the Intensive Care Unit (n=4, 10%), or home (n=8, 20%). In these latter circumstances, home was either a destroyed building or a tent.

## Discussion

This study investigates the impact of a mass casualty event (MCE) in a Palestinian hospital, focusing on the demographic characteristics of casualties, injury patterns, and hospital response within the challenging context of ongoing conflict. The findings are analysed within the framework of existing literature on conflict trauma, with emphasis on gender and age differences, injury complexity, mortality, transportation and treatment delays, and gender-specific trauma outcomes.

In this cohort, females slightly outnumbered males (51.06% *versus* 48.94%), with the age group of 18-35 years comprising the majority (55.32%) of casualties. This distribution reflects the demographic characteristics of conflict-affected civilian populations, where the indiscriminate nature of violence can affect both genders and various age groups. However, in many conflicts, men are typically at higher risk of injury because of their societal roles or active involvement in conflict zones^ix^. The relatively balanced gender distribution in our findings could suggest high exposure levels among civilians, including women and children, because of the densely populated and urbanised nature of Gaza, or perhaps even intentional targeting. The United Nations has certainly stated that women are increasingly at risk in conflict^x^.

The predominance of young adults (ages 18-35 years) aligns with findings from similar conflict settings^xi^, where younger populations are disproportionately affected because of both increased mobility and greater likelihood of being outdoors or in exposed areas. This age range is often engaged in activities outside the home, potentially placing them in harm’s way during an MCE. The high casualty rate among young adults is especially concerning, as it impacts the most productive segment of the population and has long-term socioeconomic implications.

The mean number of injuries per casualty was 1.68, reflecting the intensity and high injury burden associated with MCEs in conflict settings. This finding is consistent with research on MCEs involving high-energy trauma mechanisms, such as explosions, which typically result in multiple injuries per individual because of blast wave effects and secondary fragmentation injuries.

In non-conflict MCEs, such as those involving natural disasters or transportation accidents, the injury rate per casualty tends to be lower, as these incidents are less likely to involve high-energy blast trauma. In such events, a single blunt injury or orthopaedic trauma may be more common, rather than multiple, complex injuries. Thus, the higher injury per casualty rate in this study underscores the need for substantial medical resources, specialised trauma care, and extensive follow-up for recovery in conflict-affected hospitals.

The injuries observed in this cohort were predominantly severe and complex, including a high percentage of compound (open) injuries (78.7%) and blast-related trauma (76.6%). These patterns are consistent with conflict trauma, where blast injuries, fractures, amputations, and penetrating wounds are prevalent because of the use of explosive devices and firearms. Explosive weapons, especially in populated areas, tend to result in multi-system injuries, including fractures, amputations, and internal injuries, often necessitating advanced surgical intervention and critical care support^xii^. This injury profile significantly differs from typical civilian trauma, which often involves blunt force trauma and less frequent incidence of compound injuries.

Additionally, the range of specific injuries recorded - including fractures, amputations, and injuries to major organs – is compatible with the effects of blast injuries in an urban conflict zone. This underscores the profound impact of explosive weaponry on civilian populations and highlights the need for comprehensive trauma care resources in a zone of conflict.

The mortality rate among casualties in this study was 14.89%, with a higher mortality observed among female casualties (71.43% of deaths). Mortality rates in conflict-related MCEs can vary widely depending on factors such as injury severity, pre-hospital care, and hospital capacity. In this cohort, the high mortality rate would be compatible with delayed access to care, limited pre-hospital notification, and the complexity of injuries sustained.

In contrast, non-conflict MCEs would generally be associated with a lower mortality rate, as pre-hospital infrastructure, rapid transportation, and timely care are more accessible. Our elevated mortality rate, especially in women, may reflect systemic challenges in pre-hospital response, limited resources, and the severity of injuries sustained from explosive devices.

The mean transportation delay from the Point of Injury (POI) to hospital was 18.7 minutes, with no advance notice provided in 82.98% of cases. This delay, though relatively short considering the challenges of conflict, may still have significant consequences for patients with life-threatening injuries, particularly given the high incidence of compound trauma in this cohort. Pre-hospital notification is critical in MCEs, as it allows receiving hospitals to allocate resources, prepare trauma teams, and expedite triage.

Once in the Emergency Department (ED), the mean delay to treatment was 17.4 minutes, suggesting a fair internal hospital response despite the high patient load. We would have liked the treatment to have been quicker, but it was not possible at the time.

The disproportionately high mortality rate among female casualties in this study (71.43% of deaths) raises critical questions regarding gender-specific vulnerabilities in conflict-related trauma. While women generally constitute a smaller proportion of casualties in MCEs, studies have documented that female trauma victims often experience worse outcomes than their male counterparts^xiii^. Biological factors, such as lower mean blood volume and haemoglobin levels, may contribute to poorer outcomes among women following haemorrhagic trauma, as their capacity to withstand significant blood loss is reduced.

Furthermore, gender-based disparities in trauma outcomes may be exacerbated by sociocultural barriers that influence healthcare access, as well as differential treatment practices for women in conflict zones.

Research on gender and trauma recovery suggests that women may experience higher rates of complications, such as infection and immune response issues, following traumatic injury. These complications, coupled with the psychological impact of trauma, can exacerbate mortality risk and delay recovery in female patients. Addressing these gender-specific vulnerabilities requires a tailored approach to both pre-hospital and in-hospital care, particularly in conflict settings where the complexity of injuries necessitates specialised and individualised treatment protocols.

## Conclusion

This study underscores the profound impact of mass casualty events in conflict zones, particularly in terms of injury burden, mortality, and systemic challenges in pre-hospital and emergency care. The findings indicate that most injuries were severe, with blast-related trauma being a predominant cause of compound injuries and fatalities. The elevated mortality rate, especially among female casualties, highlights the need for gender-sensitive approaches in trauma care to address physiological and sociocultural disparities that affect outcomes.

The high rate of compound injuries and limited pre-hospital coordination point to the need for improved trauma resources and communication protocols in hospitals operating in conflict settings. Developing systems for timely pre-hospital notification, enhancing logistic support for rapid transport, and ensuring specialised trauma resources are available could mitigate the adverse effects of MCEs on civilian populations. Further research is warranted to explore the specific factors contributing to gender disparities in trauma outcomes and to develop targeted interventions that can improve survival and recovery for all affected populations.

## Data Availability

All data produced in the present study are available upon reasonable request to the authors

## Acknowledgements

Our sincerest thanks to all at the Shuhada al-Aqsa Hospital in Deir-al-Balah, Gaza, who helped make this research possible, and who were working under almost unimaginable, high-risk circumstances throughout the period of this study. Many have perished or have been displaced. Thank you also to the Palestinian Ministry of Health, to the International Rescue Committee (IRC), and Medical Aid for Palestinians (MAP).

